# A 12-year retrospective analysis of cancer epidemiology at a major oncology center in Erbil, Iraq

**DOI:** 10.64898/2026.08.05.26359839

**Authors:** Shwan Salam Marouf

## Abstract

Long-term data describing cancer patterns in Iraq remain generally limited. This retrospective observational study was conducted to evaluate the distribution and longitudinal patterns of malignant solid tumors diagnosed over a period of 12 years (2014–2025) at a major tertiary oncology center in the Kurdistan Region of Iraq. Demographic characteristics, cancer types, and temporal trend changes in cancer distribution were analyzed. Comparisons were made between the first (2014–2019) and second (2020–2025) halves of the study period. Descriptive statistics, Chi-square tests, and linear regression analyses were used to evaluate the temporal trends. After excluding records with incomplete data, 11,704 patients were included. Breast cancer was the most frequently diagnosed malignancy, accounting for over one-quarter of all cases, followed by lung, colorectal, prostate, and bladder cancers, in descending order. Women represented the majority of patients, and the mean age at diagnosis increased significantly over time. In general, the relative distribution of colorectal, genitourinary, pancreatic, uterine, and thyroid cancers increased during the study period, whereas breast and lung cancers showed a modest but significant proportional decline despite remaining the most common malignancies. A marked reduction in case numbers was observed in 2020, followed by progressive recovery in subsequent years. To conclude, cancer patterns in the Kurdistan Region of Iraq changed substantially during the 12-year study period, with increasing proportions of colorectal and several other malignancies alongside an older age at diagnosis. These findings likely reflect a combination of demographic changes, evolving lifestyle-related risk factors, and improvements in cancer detection and referral. The results provide contemporary evidence to support regional cancer control strategies, screening programs, resource allocation, and future epidemiological research.

## Introduction

Cancer represents one of the most challenging public health concerns of the 21^st^ century, associated with profound socioeconomic burdens and rising disability and death rates worldwide [1,2]. Globally, the epidemiological landscape of malignant diseases is undergoing dynamic shifts over the recent decades, driven by demographic transitions, environmental exposures, changing lifestyle situations, and advancements in preventive measures and diagnostic tools [3,4].

While comprehensive data registries in most developed countries provide robust frameworks for tracking these oncological trends, substantial gaps remain in middle- and low-income regions, making these epidemiological studies extremely necessary [5–7].

In general, accurate regional data are vital for understanding the true burden of solid tumors. In the Middle East, and specifically within the Kurdistan Region of Iraq, rapid urbanization and shifts in geopolitical and environmental determinants over the past few decades have fundamentally altered population health profiles, creating a critical need for localized, long-standing epidemiological surveillance. Variations in health infrastructure, data processing and non-centralized cancer registry systems often obscure critical shifts in disease presentation in our regions [8–11].

Furthermore, beyond overall case volumes, characterizing the vital shifts within important and high-burden malignancies—specifically breast, lung, and colorectal cancers—is critical for targeted public health strategies in our regions [12,13]. Breast cancer remains disproportionately prevalent among female populations in Middle East, frequently accounting for a staggering proportion of malignancies in women, and presenting at an earlier age and more advanced stage, compared to Western cohorts [14–16]. Similarly, changing smoking habits and environmental pollution are altering the dynamics of lung cancer, likewise shifts toward industrialized lifestyles and changing dietary traditions are implicated in the rising regional incidence of colorectal cancers [17–19].

Concurrently, critical shifts in the age distribution of cancer patients demand urgent institutional evaluation. This involves both tracking early-onset malignancies (diagnosed in individuals under the age of 50) and monitoring subtle changes in the mean age of presentation over time, alongside identifying noticeable variations in primary tumor sites across different time periods [20,21]. Mapping these temporal dynamics in oncology is vital for optimizing resource allocation for oncology centers’ infrastructure, while simultaneously tailoring public health and prevention policies to the region’s unique demographic profile [22].

The oncology registry at our tertiary care institution, being a main oncology center in the region, serves as a critical local source, capturing a diverse patient population over an extended geographical area. Retrospective analyses of such large-scale institutional databases offer invaluable insights into the shifting dynamics of cancer presentation, bypassing the limitations of short-term cross-sectional evaluations. By analyzing clinical trends across a sustained timeframe, it becomes possible to distinguish transient annual fluctuations from factual, long-term epidemiological shifts in cancer presentations. To address these critical data gaps, this study presents a comprehensive 12-year retrospective epidemiological analysis of malignant solid tumors in patients aged over 14 years, registered at our center between January 2014 and December 2025.

The primary objectives of this study were to characterize the distribution of solid malignant tumors stratified by primary disease site, gender, and age in our region; and secondly, to evaluate temporal trends in time by analyzing annual diagnostic patterns and detecting important shifts between two consecutive six-year periods (2014–2019 versus 2020–2025). Ultimately, these findings, together with further broader epidemiological studies, aim to provide a robust and evidence-based foundation to enlighten clinical practice, guide regional oncology policy, and contribute to the broader mapping of cancer epidemiology in the Kurdistan Region of Iraq.

## Methods

### Study Design

This retrospective observational study was conducted at Rizgary Oncology Center in Erbil, Iraq, a major tertiary cancer referral center serving the Kurdistan Region and surrounding areas. It evaluated demographic trends among patients diagnosed with malignant tumors over a 12-year period from January 2014 to December 2025.

### Study Population and Inclusion Criteria

All patients (≥14 years), who presented with newly diagnosed malignant solid tumors and registered at the center during the study period (January 2014 to December 2025), were included. To assemble the study cohort, electronic health records of eligible cancer patients were extracted from the center’s registry database based on standardized International Classification of Diseases, Tenth Revision (ICD-10) site-specific codes. To be included in the study, patients were required to have a histopathologically confirmed diagnosis of a solid malignancy in a known/trusted histopathology laboratory. Complete baseline records containing essential demographic and diagnostic parameters—specifically age, gender, and year of diagnosis—were required for final eligibility.

### Exclusion Criteria

Patients were excluded if they presented with benign tumors or hematological malignancies. Additionally, duplicate records and cases with missing or incomplete essential medical and demographic data were omitted from the study.

### Data Collection

Demographic and diagnostic data of the patients were extracted from an institutional electronic database managed by the center’s trained medical staff. This database serves as a digital index of the primary paper-based patient charts/case-notes, capturing key baseline characteristics—such as age, gender, cancer subtype, and year of diagnosis—immediately upon initial presentation to the center. Relevant data points were compiled using a standardized collection form. To maintain patient confidentiality, all personal identifiers, including patient names and contact details, were completely removed from the dataset prior to data analysis. Data were accessed for the research purpose on 22 June 2026. Because all patient records were fully anonymized prior to statistical evaluation, the author had no access to information that could identify individual participants during or after data collection.

### Age Classification

In addition to analyzing age at diagnosis as a continuous variable to calculate the mean and other descriptive statistics, age at diagnosis was categorized into five predefined groups: <40, 40–49, 50–59, 60–69 and ≥70 years. These categories were selected to allow evaluation of early-onset malignancies and age-related changes in cancer presentation over time.

### Cancer Type Classification

In addition to separate assessment for individual cancer types based on IDC-10 codes, selected analyses were performed for aggregated cancer groups based on their major histological/anatomical sites of origin (e.g. sarcomas, central nervous system, gastrointestinal, genitourinary, gynecologic and thoracic cancers). The “Other/Rare Tumors” category in the cancer grouping comprised infrequently encountered malignancies, for example neuroendocrine tumors and tumors originating from rare anatomical locations such as mucosal melanomas, that were not sufficiently signified to warrant a separate analysis.

### Time Stratification

In addition to examining the year of diagnosis annually from 2014 to 2025, the 12-year study timeframe was stratified into two consecutive 6-year intervals to evaluate broader temporal trends. The first half of the study (Period-1) spanned from 2014 to 2019, and the second half (Period-2) encompassed 2020 through 2025. This stratification allowed assessment of demographic changes across time while maintaining adequate sample size within each study period.

### Statistical Analysis

Descriptive statistics were used to summarize patient demographics and cancer distribution. Categorical variables, such as age group, gender and cancer type, were presented as frequencies and percentages.

Baseline characteristics and cancer distributions were compared between Period-1 (2014– 2019) and Period-2 (2020–2025). The percentage difference in distribution between the two periods was calculated for each cancer type. Proportional differences between the two study periods were analyzed using the Chi-square test. A p-value of less than 0.05 was considered statistically significant.

To assess chronological trends in the age at diagnosis over the 12-year study interval, simple linear regression analysis was performed. In this model, calendar year was entered as a continuous independent predictor, and age-at-diagnosis served as the dependent variable.

Trends were evaluated for the aggregate cancer cohort as well as individually for specific common malignancies, namely breast, lung, and colorectal cancers.

Data curation, percentage change calculations, and preliminary descriptive statistics were managed using Microsoft Excel (Version 2024; Microsoft Corp., Redmond, WA, USA). Formal inferential statistical testing, including Chi-square analysis, was conducted using JASP (Version 0.97.1; JASP Team, Amsterdam, Netherlands).

### Ethical Considerations

This study was approved by the Scientific Research Ethical Committee of Hawler Medical University in Erbil, Iraq (Ref: HMUD/2527, approved June 7, 2026) and complied with all required ethical standards. Prior to initiation, the study protocol was presented as a seminar and approved by the institutional scientific committee. Because this project involved a retrospective analysis of institutional data, no direct patient contact occurred. To safeguard participant confidentiality, all identifying information was removed from the dataset before data analysis began.

## Results

Initially a total of 12,046 patient records were identified and reviewed during the study period from 2014 to 2025. Of this primary cohort, 342 records were excluded because of incomplete or missing essential data, leaving 11,704 patients diagnosed with malignant solid tumors eligible for inclusion in the final analysis. Males constituted 43.6%, while females represented 56.4% of the studied population. The median age at diagnosis was 57 years, ranging from 14 to 111 years. While the largest age group was patients in their 60s (accounting for greater than 23% of cases), the study population demonstrates a prominent concentration of cases in the relatively younger and middle-aged individuals, with patients under the age of 50 account for over one-third of the entire cohort (Table 1).

**Table 1.** Baseline demographic characteristics of the study population (2014–2025)

| Characteristic | Total cohort (N = 11,704) |
| --- | --- |
| Gender |  |
| Male | 5,108 (43.6%) |
| Female | 6,596 (56.4%) |
| Age at diagnosis (years) |  |
| Median age | 57 |
| Mean age ± SD | 56.02 ± 15.77 |
| Age range | 14–111 |
| Age group distribution (years) |  |
| <40 | 1,855 (15.9%) |
| 40–49 | 2,114 (18.1%) |
| 50–59 | 2,452 (20.9%) |
| 60–69 | 2,762 (23.6%) |
| $\geq 70$ | 2,521 (21.5%) |

Table 2 presents the annual distribution of the total number of cancer cases diagnosed between 2014 and 2025. The number of recorded cases varied over the study period; the highest annual case counts were observed in the years 2014 (1,192 cases; 10.2%) and 2015 (1,195 cases; 10.2%). This was followed by a gradual decline that reached its lowest level in 2020 (674 cases; 5.8%). Thereafter, the number of cases increased steadily again, reaching 1,110 cases (9.5%) in 2025. In general, the annual number of recorded cancer cases demonstrated temporal yearly fluctuations, with a marked reduction during the middle years of the study period followed by a progressive recovery in the subsequent years.

**Table 2.** Annual number of recorded cancer cases (2014 – 2025)

| **Year of presentation** | **Total cancer cases** |
| --- | --- |
| 2014 | 1,192 (10.2%) |
| 2015 | 1,195 (10.2%) |
| 2016 | 965 (8.2%) |
| 2017 | 814 (7.0%) |
| 2018 | 900 (7.7%) |
| 2019 | 861 (7.4%) |
| 2020 | 674 (5.8%) |
| 2021 | 893 (7.6%) |
| 2022 | 1,028 (8.8 %) |
| 2023 | 1,002 (8.6%) |
| 2024 | 1,070 (9.0%) |
| 2025 | 1,110 (9.5%) |

The aggregate distribution of all documented cancer groups across the entire study cohort is outlined in Table 3. As shown, breast cancer was the most frequently diagnosed malignancy, accounting for more than a quarter of all diagnosed cases. Genitourinary cancers (including prostate, urinary bladder, kidney and testicular cancers combined) occupied the second place in the table at about 15%. Thoracic, colorectal, gynecological and upper gastrointestinal malignancies were followed in a descending order of frequency. Intermediate frequencies were observed for head and neck cancers, central nervous system tumors and sarcomas. Conversely, endocrine, pancreatic, hepatobiliary and skin cancers were much less common, each accounting for only a small fraction of the total cohort.

**Table 3.** Overall distribution of major cancer groups among the study population.

| Cancer type | Number | Percentage |
| --- | --- | --- |
| Breast | 3,223 | 27.54% |
| Genitourinary | 1,754 | 14.99% |
| Thoracic | 1,335 | 11.40% |
| Colorectal | 1,170 | 10.00% |
| Gynecologic | 809 | 6.90% |
| Upper Gastrointestinal | 617 | 5.27% |
| Head and Neck | 495 | 4.23% |
| Central Nervous System | 469 | 4.01% |
| Sarcoma (Soft Tissue and Bone) | 428 | 3.66% |
| Endocrine | 367 | 3.14% |
| Pancreas | 285 | 2.44% |
| Hepatobiliary | 237 | 2.02% |
| Skin | 209 | 1.79% |
| Unknown Primary | 219 | 1.87% |
| Other/Rare Tumors | 87 | 0.74% |
| Total | 11,704 | 100.0% |

Based on the results of the present study, among male patients, as seen in Fig 1, lung cancer was the most frequently diagnosed malignancy, accounting for 19.6% of cases, followed by prostate cancer at15.5%, colorectal cancer at 11.4% and urinary bladder cancer at 7.8%.

**Figure 1.**
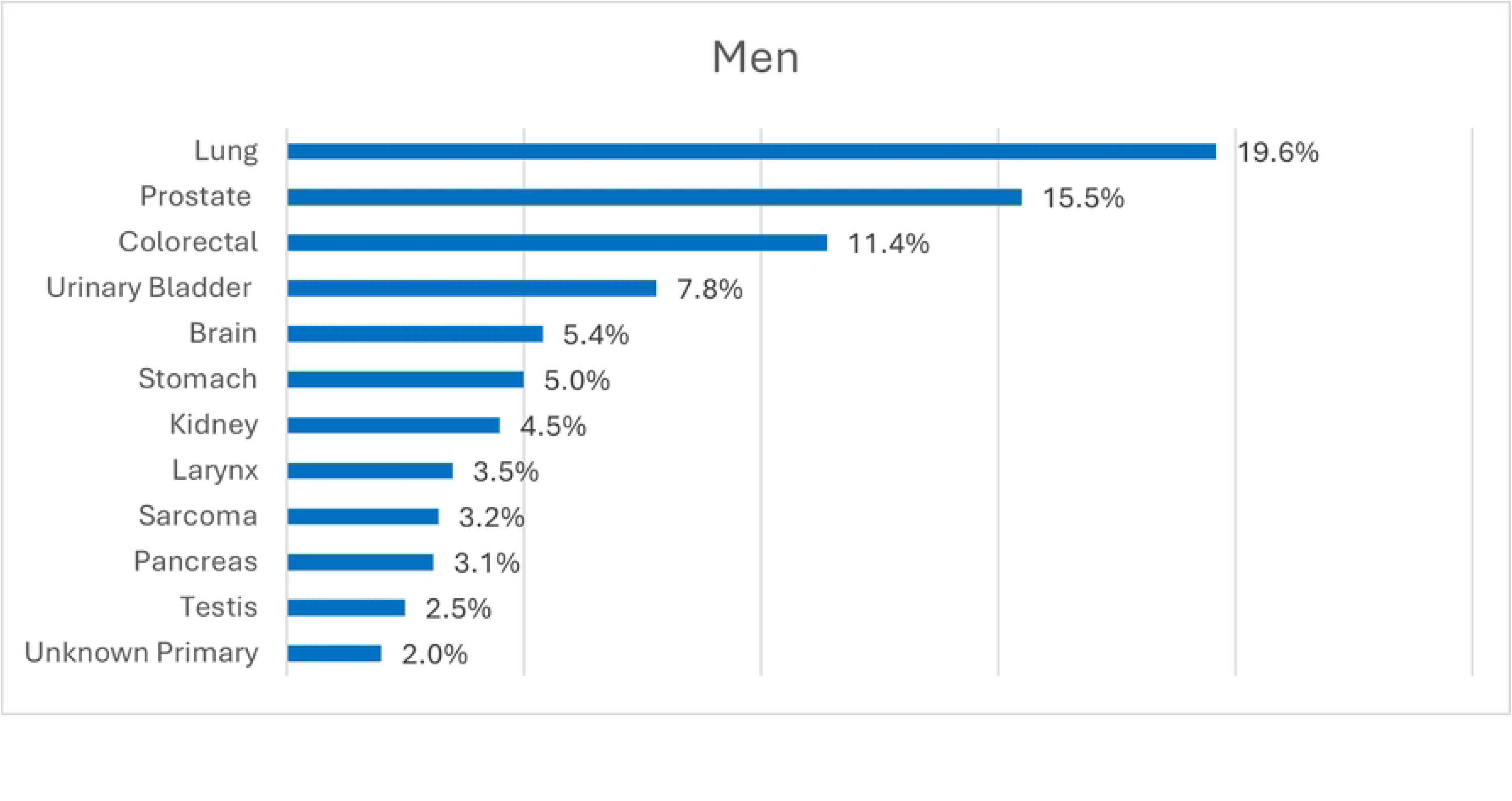
Cancer types distribution among male patients. The bar charts represent the percentage distribution of cancer types in men during the study period.

Meanwhile, sarcomas, pancreatic and testicular cancers were among the least frequent malignancies diagnosed in men at around 2 – 3% for each type. Regarding female patients (Fig 2) on the other hand, breast cancer was by far the most predominant malignancy, representing 47.6% of all solid cancerous tumors diagnosed in women; this was followed by colorectal, ovarian, lung, uterine and thyroid cancers at 7.6%, 5.0%, 4.7%, 4.6% and 4.1% respectively. These findings demonstrate distinct gender-specific patterns in cancer distribution, with lung, prostate and urinary bladder cancers predominating in men, whereas breast cancer constituted the major cancer burden among women accounting for nearly half of the female cohort.

**Figure 2.**
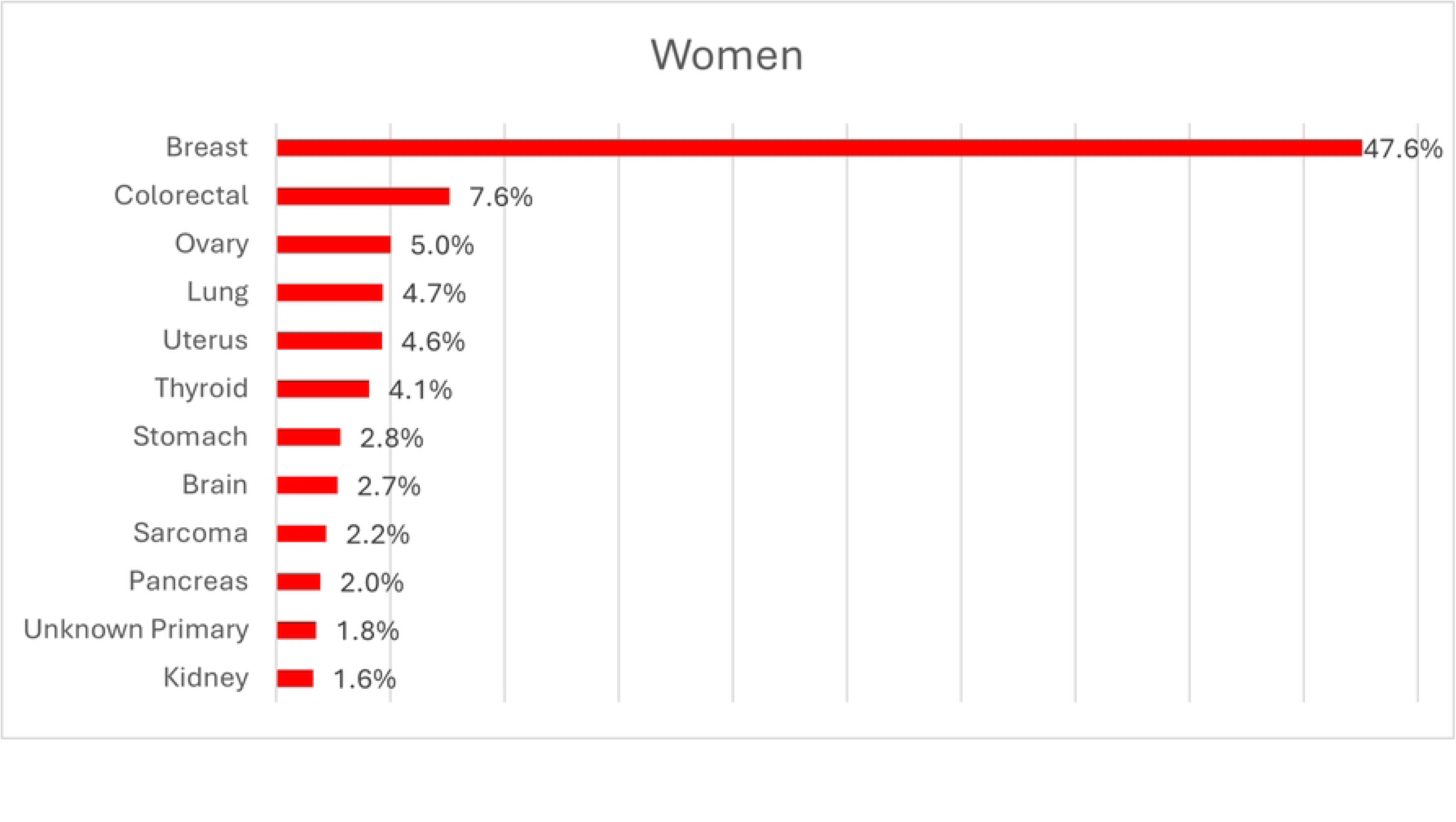
Cancer types distribution among female patients. The bar charts represent the percentage distribution of cancer types in women during the study period.

The general distribution of patients’ age at the time of cancer diagnosis across the whole study period (2014–2025) is illustrated in Fig 3. Although the age distribution remained broadly similar throughout the study years, a gradual increase in the median age at cancer diagnosis was observed over time. Analysis of variance demonstrated a statistically significant difference in mean age among the study years (ANOVA, *P* < 0.001). Besides, linear regression analysis identified a significant positive association between calendar year and age at diagnosis of cancer in general (unstandardized coefficient = 0.272, *P* < 0.001), indicating an average increase of approximately 0.27 years in patient’s age at diagnosis for each successive year of the study period. Despite this statistically significant trend, the magnitude of the increase was relatively small, with considerable overlap in the age distributions across the study years.

**Figure 3.**
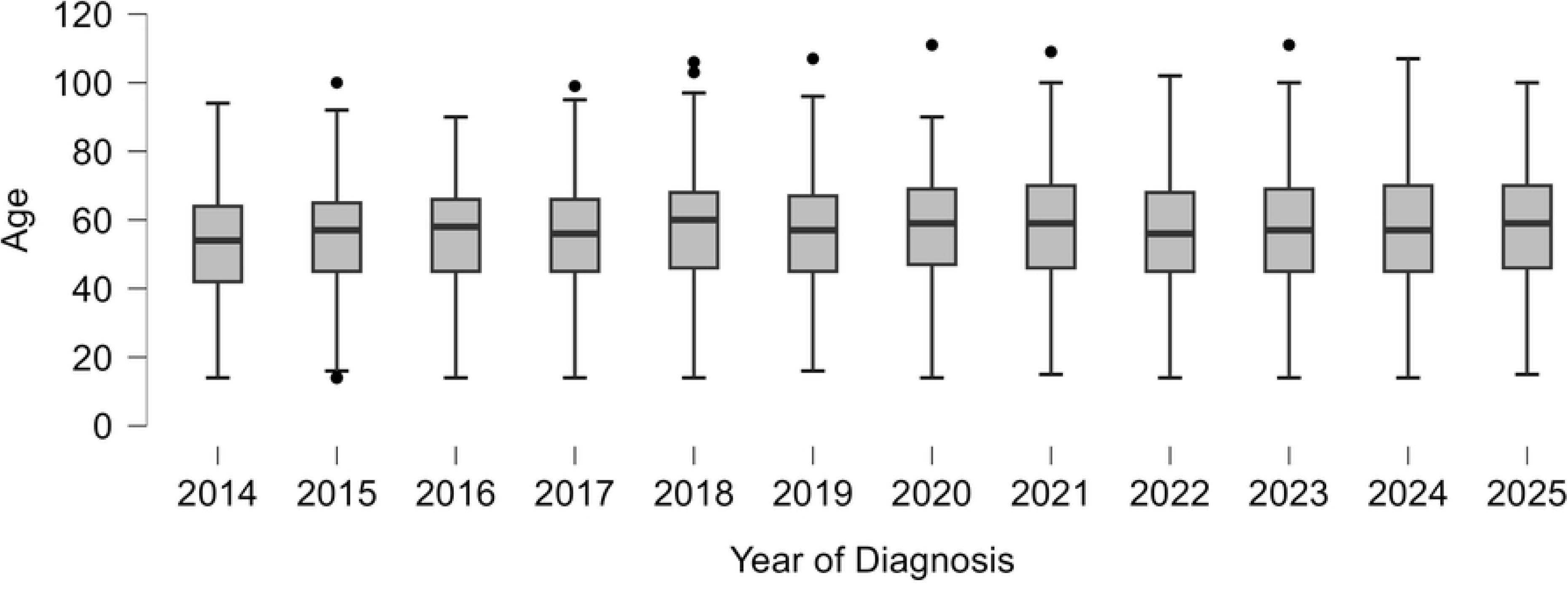
Annual age distribution at diagnosis from 2014 to 2025. Box plots illustrate the median (solid horizontal line), interquartile range (boxes), minimum/maximum values (whiskers), and outliers (dots) for each year.

To further explore whether this pattern was consistent across different types of malignancies, age distributions over time for the most common cancer types, including breast, lung and colorectal cancers were studied separately (Fig 4) to show comparison of age-related trends across the study period.

**Figure 4.**
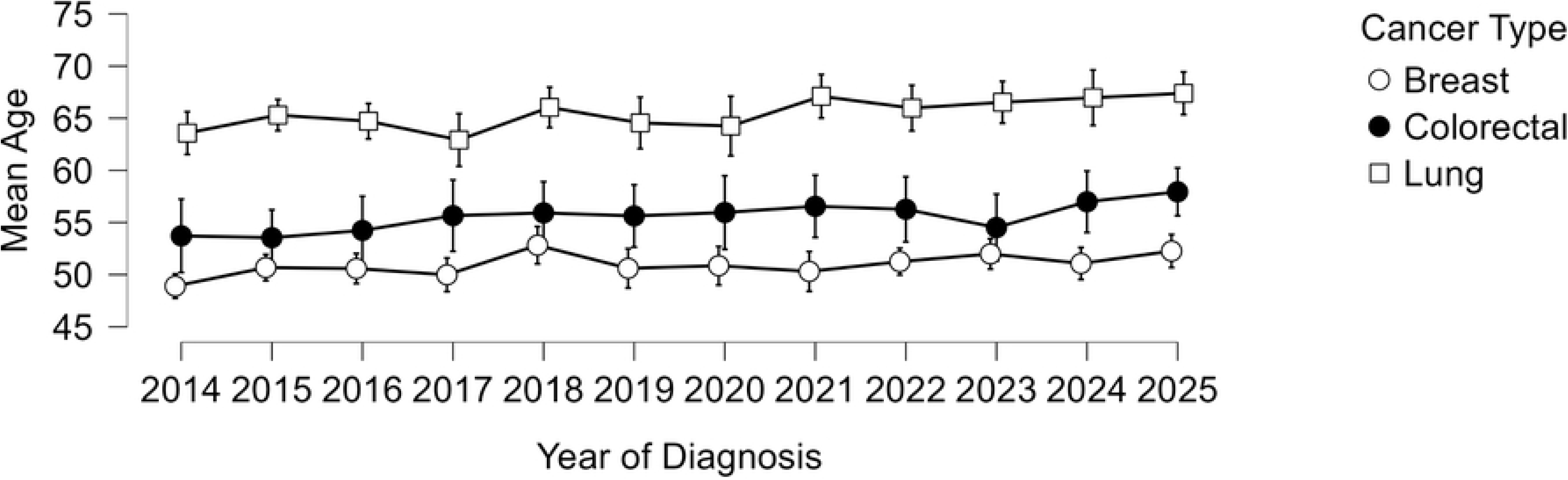
Comparison of annual mean age trends for breast (open circle), colorectal (solid circle), and lung cancer (open square). Error bars indicate 95% confidence intervals.

For breast cancer, the age at diagnosis changed significantly across the study years. Linear regression analysis demonstrated a significant increase in median age at diagnosis of approximately 0.195 years per calendar year (unstandardized coefficient = 0.195, *P* = 0.001).

Similarly, among patients with lung cancer, linear regression identified a significant positive association between calendar year and age at diagnosis with an estimated rise by an average of 0.284 years per calendar year (unstandardized coefficient = 0.284, *P* = 0.001).

Moreover, a significant positive linear trend was observed regarding the patient’s age at the time of colorectal cancer detection; the average age at diagnosis advanced by 0.312 years per calendar year (unstandardized coefficient = 0.312, *P* = 0.011).

Overall, these findings suggest a gradual but modest increase in the mean age at cancer diagnosis over the course of the study years, both for the total cancer population and for the common malignancies, especially breast, lung and colorectal cancers. Thus, contrary to concerns regarding a shift toward younger age at cancer diagnosis, the present study demonstrated a gradual increase in the age at diagnosis, with a significant yet modest positive trend for breast, lung, and colorectal cancers.

Furthermore, Table 4 summarizes the trends in the distribution of individual cancer types diagnosed across the two study periods—comparing the first half (2014–2019) with the second half of the study (2020–2025). Breast cancer remained the most frequently diagnosed malignancy in both periods; however, its relative proportion showed a statistically significant decline from 28.8% in the first half to 26.2% in the second study half (p = 0.002). Similarly, a significant reduction was observed in the lung cancer cases, which decreased from 12.3% to 10.1% (p < 0.001).

**Table 4.**
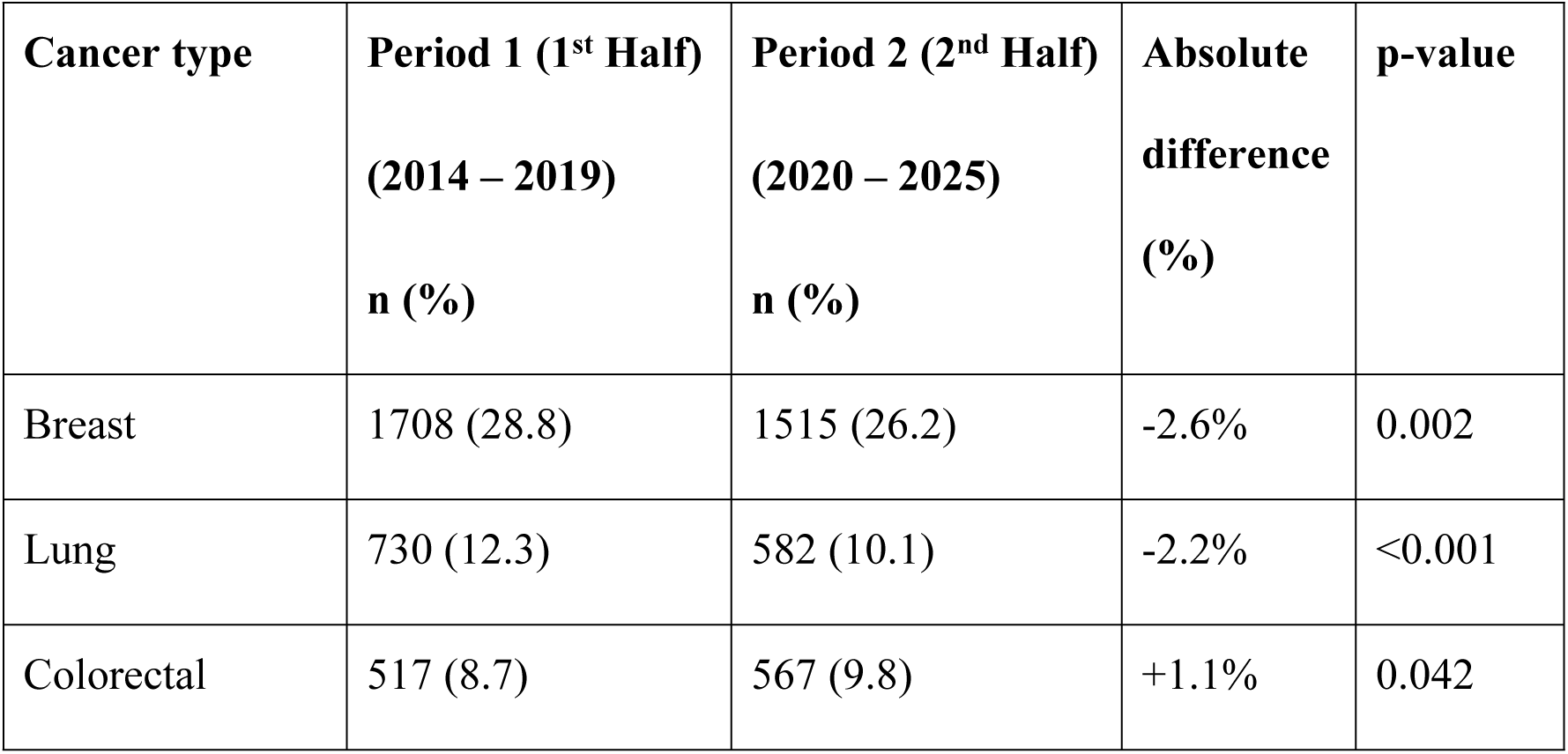
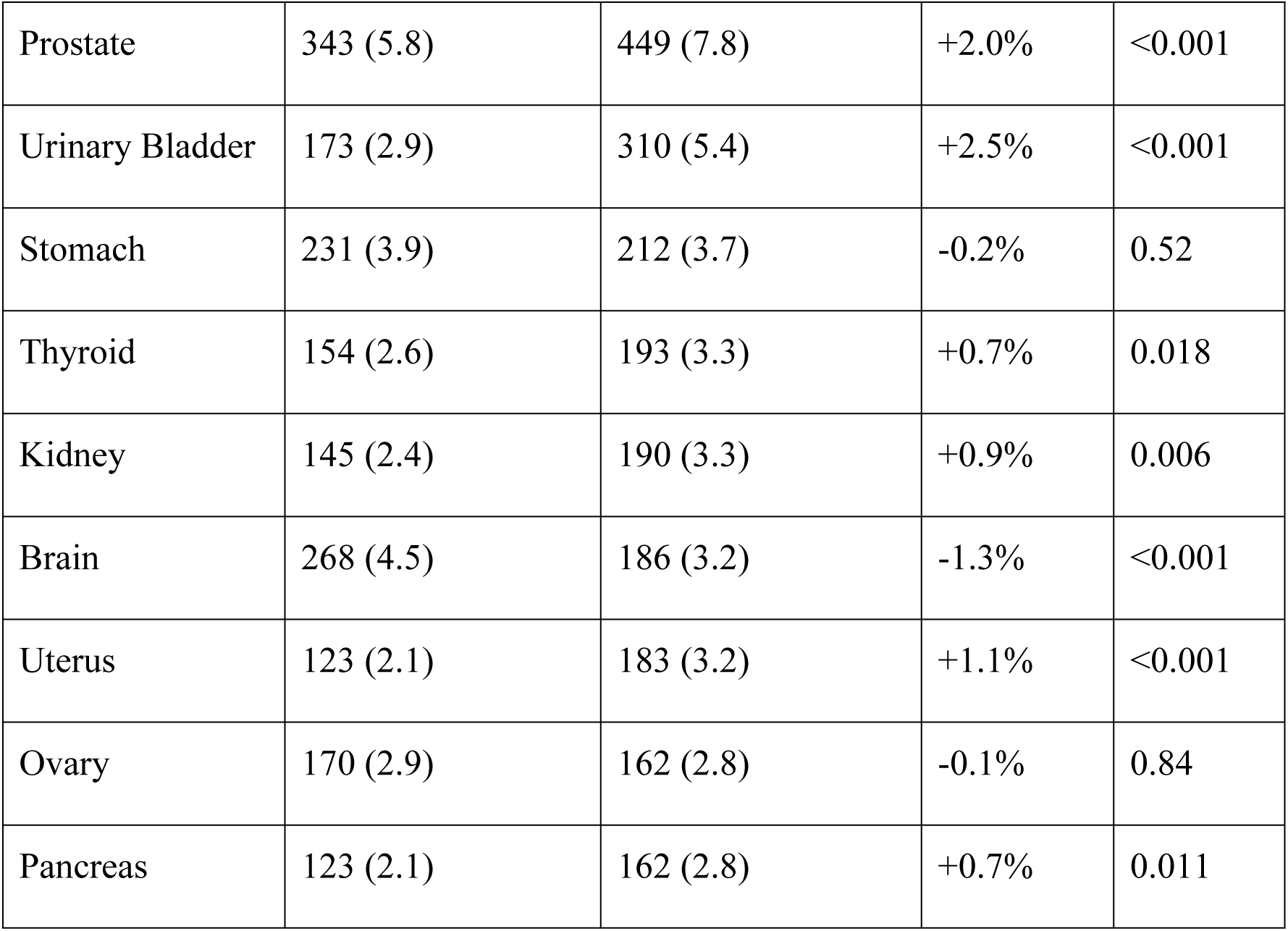
Distribution and absolute difference of major individual cancer types across the two study periods.

| Cancer type | Period 1 (1<sup>st</sup> Half)<br>(2014 – 2019)<br>n (%) | Period 2 (2<sup>nd</sup> Half)<br>(2020 – 2025)<br>n (%) | Absolute<br>difference<br>(%) | p-value |
| --- | --- | --- | --- | --- |
| Breast | 1708 (28.8) | 1515 (26.2) | -2.6% | 0.002 |
| Lung | 730 (12.3) | 582 (10.1) | -2.2% | <0.001 |
| Colorectal | 517 (8.7) | 567 (9.8) | +1.1% | 0.042 |
| Prostate | 343 (5.8) | 449 (7.8) | +2.0% | <0.001 |
| Urinary Bladder | 173 (2.9) | 310 (5.4) | +2.5% | <0.001 |
| Stomach | 231 (3.9) | 212 (3.7) | -0.2% | 0.52 |
| Thyroid | 154 (2.6) | 193 (3.3) | +0.7% | 0.018 |
| Kidney | 145 (2.4) | 190 (3.3) | +0.9% | 0.006 |
| Brain | 268 (4.5) | 186 (3.2) | -1.3% | <0.001 |
| Uterus | 123 (2.1) | 183 (3.2) | +1.1% | <0.001 |
| Ovary | 170 (2.9) | 162 (2.8) | -0.1% | 0.84 |
| Pancreas | 123 (2.1) | 162 (2.8) | +0.7% | 0.011 |

In contrast, an increasing trend was observed for several other cancer types. For instance, colorectal, prostate and urinary bladder cancers demonstrated a statistically significant rise in their relative frequencies between the two study halves. As a whole, significant temporal changes were observed in the distribution of several cancer types, whereas others remained relatively stable across the two halves of the study period. These findings indicate a shift in the distribution pattern of some major cancer types over time, characterized by a relative decline in breast, lung and brain tumors with a corresponding increase in genitourinary, colorectal, pancreatic, uterine and thyroid cancers.

## Discussion

The study provides a comprehensive overview of cancer epidemiology over a 12-year period in Kurdistan Region of Iraq. In general, females accounted for a greater proportion of cancer cases than males (56.4% vs. 43.6%), a finding that is largely consistent with several cancer reports from Iraqi Cancer Registry and previous Iraqi cancer care studies [23,24], where the high incidence of breast cancer combined with gynecological malignancies contributes substantially to the overall cancer burden in women. Regionally, this gender distribution aligns with data from the Global Cancer Observatory, which notes similar trends across neighboring countries [25].

Although global registries often note a higher overall cancer occurrence in men due to worldwide high rates of lung, prostate, and colorectal cancers, our findings reflect the unique demographic and epidemiological shift captured in the World Health Organization’s developing country profiles where the solid tumor burden inclines significantly toward the female population [26].

The median age of the patients at the time of cancer diagnosis in the present study was 57 years, which is comparable to findings from Iraq and several other Middle Eastern countries, where the average age of patients at cancer diagnosis generally ranges between the mid-fifties and early sixties [27]. This relatively younger age at diagnosis compared with the well-developed countries, where the peak incidence of solid tumors shifts significantly into later decades of life [1], reflects the younger population structure of our region, differences in life-style characteristics, and a higher contribution of cancers that occur at younger ages, particularly breast, colorectal and thyroid cancers. [25,28]

The considerable burden of cancer in individuals under 50 years observed in our cohort emphasizes the need for greater awareness and better preventive, diagnostic and therapeutic strategies in younger populations in our region.

A notable finding in the present study was the gradual decline in the total number of registered cancer cases during the middle years of the study period, particularly around 2020– 2021. This temporal registration decline likely reflects the indirect effects of the COVID-19 pandemic rather than a true reduction in cancer incidence. Worldwide, the pandemic disrupted healthcare services, defective transport and reduced access to diagnostic and therapeutic facilities, leading to substantial reductions in newly diagnosed cancer cases. A large systematic review and meta-analysis reported about 27% decrease in cancer diagnoses during the pandemic, with the greatest reductions observed during the early months of 2020, followed by a gradual recovery [29]. Similar declines in cancer registrations have been reported across multiple countries and cancer registries, supporting the interpretation that the observed reduction in the records of newly diagnosed cases in the present study was largely attributable to pandemic-related healthcare disruptions rather than true changes in cancer occurrence.[30]

Notably, during the study period between 2014 and 2025, there was a steady upward shift in the average age of the patients at the time of cancer diagnosis. This observation is consistent with international demographic trends and cancer surveillance data, which indicate that population aging is a major driver of cancer burden globally [2,31,32]. Similar age-related shifts have been reported in several regional studies across the Middle East, reflecting changes in population structure and increase longevity [33,34].

The accumulation of age-related risk factors and enhanced diagnostic capabilities may contribute to the higher detection of cancers among relatively older patients who might previously have remained undiagnosed. These findings have important implications in healthcare planning, as older patients often require more sophisticated and multidisciplinary management strategies.

Throughout the 12 years duration of the study, breast and lung cancers remained the most prevalent malignancies in women and men, respectively. Notably, however, the data revealed a modest but yet statistically significant decrease in the relative proportion of both cancer types over time. This finding contrasts with numerous international reports, where the incidence of both breast and lung cancers have generally continued to increase despite improvements in prevention and treatment; this is largely driven by population ageing, urbanization, lifestyle changes, and improved cancer detection [1,2,35].

This apparent decline observed in the present study should therefore be interpreted cautiously. Because this analysis evaluated the relative distribution of cancers registered at a single regional tertiary oncology center rather than population-based age-standardized incidence rates, a reduction in the proportion of breast and lung cancers does not necessarily indicate a true decline in their incidence. Instead, it may reflect the substantially faster increases observed in several other malignancy types, particularly colorectal, prostate, urinary bladder, kidney, thyroid, pancreatic, and uterine cancers, which consequently reduced the proportional contribution of breast and lung cancers to the overall cancer burden. Recently, several additional factors such as improvements in diagnostic capacity and referral pathways may have preferentially increased the detection rates of cancers that were previously underdiagnosed, particularly gastrointestinal and genitourinary malignancies.

The increasing proportions of colorectal and genitourinary cancers observed over the latter half of the study period are consistent with the epidemiological transition reported across many developing and middle-income countries [24,36].

The rise in colorectal cancer has been widely attributed to population aging, and lifestyle-related risk factors such as dietary westernization, increasing obesity and physical inactivity that have become increasingly prevalent in Iraq and other Middle East and North African countries [35,37].

Similarly, the increasing proportion of genitourinary cancers, especially of prostate and urinary bladder in men, probably reflects both a true increase in disease incidence, likely due to population aging and smoking, and an improved case detection rate through greater clinical awareness and more widespread use of diagnostic procedures such as cystoscopy, prostate-specific antigen (PSA) and transrectal ultrasound. [38,39]

The modest increases in pancreatic and uterine cancers are also consistent with global trends and may be driven by increasing prevalence of obesity, insulin resistance, and aging populations, all of which are well-established risk factors for such cancers [40–42].

In addition, the slight rise in thyroid cancer cases with time reflects observations from several regional and international studies and is likely partly attributable to increased use of high-resolution ultrasonic imaging and guided needle aspirations/biopsies that improved detection of small or indolent tumors, although a genuine increase in incidence cannot be excluded [43].

Altogether, these findings suggest that the cancer profile in the Kurdistan Region of Iraq is progressively shifting toward malignancies associated with aging, metabolic disorders, and lifestyle-related exposures, paralleling patterns reported in other countries undergoing similar demographic and socioeconomic transition.

## Conclusions

This 12-year population-based analysis demonstrated a substantial and evolving cancer burden in the Kurdistan Region of Iraq. Breast cancer remained the most common malignancy overall, while notable increases were observed in colorectal, prostate, urinary bladder, kidney, thyroid, pancreatic, and uterine cancers. Additionally, the age at cancer diagnosis showed a significant upward trend during the study period.

These findings likely reflect the combined effects of demographic aging, urbanization, lifestyle modifications, environmental influences, and improvements in cancer detection and registration systems. The observed epidemiological transition underscores the need for strengthened cancer prevention strategies, expansion of screening programs, earlier detection initiatives, and enhanced healthcare resource allocation.

Future multicenter studies incorporating survival outcomes, risk factor assessment, and molecular characteristics are warranted to further clarify the evolving cancer landscape in the region and to guide evidence-based cancer control policies.

## Data Availability

All relevant data are within the manuscript and its Supporting Information files.

## Acknowledgments

I extend my sincere appreciation to the leadership and staff of the Oncology Center at Rizgary Hospital for granting access to the institutional registry files and for their administrative assistance during data retrieval. Special gratitude is also extended to my family for their unwavering encouragement, patience, and support throughout the preparation of this manuscript.

## Notes

### Competing Interest Statement

The authors have declared no competing interest.

### Author Declarations

This study was approved by the Scientific Research Ethical Committee of Hawler Medical University in Erbil, Iraq (Ref: HMUD/2527, approved June 7, 2026) and complied with all required ethical standards.

